# Long-term Effects of Text Messages with Financial Incentives for Men with Obesity: Two-year Follow-up of the Game of Stones trial

**DOI:** 10.1101/2024.12.12.24318921

**Authors:** Stephan U Dombrowski, Pat Hoddinott, James Swingler, Lisa Macaulay, Catriona O’Dolan, Seonaidh Cotton, Alison Avenell, Abraham M Getaneh, Cindy M Gray, Kate Hunt, Frank Kee, Alice MacLean, Michelle C McKinley, Claire Torrens, Katrina Turner, Marjon van der Pol, Graeme MacLennan

## Abstract

**Objectives:** To compare the difference in percentage weight change at 24 months following a 12-month text messaging with financial incentive intervention compared to a waiting list control group, and to compare the text messaging alone group to the control group.

**Design:** Assessor-blinded three-arm randomised clinical trial.

**Setting:** Three disadvantaged communities from the UK.

**Participants:** 585 men with body mass index (BMI) of ≥30kg/m^2^ enrolled between July 2021 and May 2022, of which 377 participants (64%) completed 24 months’ follow-up.

**Interventions:** 12 months of daily behavioural text messages combined with financial incentives; 12 months of the same text messages alone; or waiting for 12 months followed by receipt of the first 3 months (months 12-15) of text messages (control).

**Main outcome measures:** Within-participant change from baseline weight.

**Results:** At 24 months, the mean percent weight change (standard deviation) was −3.9% (6.9%) for the text messaging with financial incentives group, −2.6% (6.8%) for the text messaging alone group, and −2.2% (6.8%) for the control group. Compared with the control group, the mean percent weight change was not significantly greater in the text messaging with financial incentives group (mean difference, −1.0%; 97.5% CI, −2.6 to 0.6; P = .22) or the text messaging alone group (mean difference, −0.0%; 97.5% CI, −1.6 to 1.5; P = .95). At least 5% weight loss at 24 months was achieved by 52 (40%) participants in the text messaging with financial incentives group; 32 (28%) in the text messaging alone group and 43 (32%) in the control group.

**Conclusions:** A scalable, low-cost text message with financial incentives intervention supported clinically relevant maintenance of weight loss 12 months after the intervention ceased, including those from disadvantaged backgrounds.

**What is already known on this topic:**

- Men, particularly those in lower socio-economic groups, engage less frequently in existing weight loss interventions and services, and might benefit from gender-sensitized interventions.
- The Game of Stones trial showed that a 12-month intervention consisting of text messaging with endowment financial incentives resulted in statistically significant weight loss in men with obesity compared with control.

**What this study adds:**

- At 24 months, participants receiving 12 months of behavioural text messages with financial incentives displayed continued weight loss which is likely to be clinically important for some men.
- This digital, scalable, low risk text messaging with financial incentives intervention, with low participant and staff burden, shows promise for longer-term weight maintenance in men, including those living in the most deprived areas.

## Background

Obesity increases the risk of type 2 diabetes, heart disease, stroke, mobility problems and some cancers, and its prevalence is high and rising^1^. In 2021 to 2022, the prevalence of adults living with obesity remained similar among women (26.1%) and men (25.8%)^2^. However, men engage less in existing weight loss interventions and services, suggesting that they might benefit from gender-sensitized interventions^3^.

Behavioural interventions remain a cornerstone of obesity treatment and have the potential to support weight change in the short and longer term^4^ ^5^, however there is an evidence gap for engaging men in lower socio-economic groups^6^. Long-term outcomes of scalable behaviour change interventions for weight loss are required to inform cost-effectiveness. A systematic review of 15 randomised controlled trials of text message-delivered weight management interventions found no studies with follow up beyond 12 months, and none focused on men^7^. In a systematic review of financial incentives for weight loss including 35 randomised controlled trials, the median intervention duration was 3 months, 15 trials followed-up post intervention, with 10.5 months being the longest follow-up^8^.

Game of Stones is a 12 month three-arm pragmatic randomised controlled trial of 585 men with obesity who received daily behavioural text messaging over 12 months, with and without financial incentives related to weight-loss targets, compared to a waiting list control group who received 3 months of text after primary outcome measurement at 12 months. The protocol^9^ and the 12 month results are published^10^. Underserved groups were recruited with 39% (n=227) living in the more disadvantaged areas, 40% (n=235) with multiple long-term conditions, 29% (n=165) with a physical or mental disability and 25% (n=146) ever diagnosed with a mental health condition. The 12 month findings showed that text messaging with financial incentives significantly improved weight loss compared to the waiting list control group (mean difference, −3.2%; 97.5% CI, −4.6 to −1.9; P < .001), whereas text messaging alone was not significantly better than the control (mean difference, −1.4%; 97.5%CI, −2.9 to 0.0, P = .05)^10^.

The two primary objectives for this paper are to compare the difference in percentage weight change at 24 months for the 12-month text messaging with financial incentives group compared to the waiting list control group (who received texts for 3 months between months 12-15), and to compare the text messaging alone group to the waiting list control group. The secondary objectives were to examine differences between groups in other weight outcomes, to compare PHQ-4, Quality of Life (EQ-5D-5L), EQ-5D-5L Anxiety and Depression (AD) dimension measures, and to examine differences in exploratory outcomes of weight management strategies used, satisfaction with weight loss progress and propensity to recommend of Game of Stones to others.

## Methods

### Trial design

Game of Stones was conducted between July 2021 to July 2023 in three UK areas with disadvantaged communities: Belfast, Bristol and Glasgow. The sample size was pre-specified for 12 months only. Overall, 585 men were recruited with a body mass index (BMI) ≥30kg/m^2^. Men were recruited either through general practices including letters and posters, or via community recruitment including information stands in supermarkets, posters, social media, and word of mouth.

The three study groups were: i) 12 months of text messaging with financial incentives. Daily automated behavioural text messages based on psychological theory embedding evidence-based behaviour change techniques designed to support weight management. Loss-framed incentives were offered in which money was ‘lost’ from an initial endowment of £400 by not meeting verified weight loss targets in comparison with baseline weight (5% loss at 3 months, 10% loss at 6 months and maintaining 10% loss weight loss at 12 months); ii) 12 months of the same behavioural text messages alone; or iii) a 12-month waiting list followed by the first three months of the intervention groups’ text messages between months 12 and 15, followed by 9 months of no intervention.

### Measures and outcomes

The primary outcome for this analysis was within-participant change from baseline weight expressed as a percentage of baseline weight at 24 months. Staff measuring weight and analysing outcomes were unaware of group allocations. Weight was measured in-person within 23 days of the target date for follow-up using study scales and verified independently by another researcher. If the participant was not able to return for measurement of weight in person, participants were offered a video call to measure weight on study scales delivered to their home. If participants declined or did not respond to this option, they were mailed a letter requesting email or postal return of a questionnaire with self-reported weight on their own scale.

The secondary outcomes were: absolute weight change from baseline (kg); weight loss categories (including: no weight loss); >0% weight loss; ≥5% weight loss; ≥ 10% weight loss; quality of life using the EQ-5D-5L overall utility score; EQ-5D-5L visual analogue scale; EQ-5D-5L Anxiety and Depression dimension^11^; mental health assessed using the four questions of the Patient Health Questionnaire-4 (PHQ-4)^12^.

The exploratory outcomes at 24 months were: weight management strategies assessed by asking, “Which of these strategies have you used in the last 12 months to lose weight?” together with 13 response options (e.g. “Had a weight goal to work towards”) based on evidence of effective strategies for weight management^13^; satisfaction with weight loss progress, assessed by asking “Given the effort you put into losing weight, how happy are you with your progress at the moment?” with responses on a 7-point scale ranging from 1 (very unhappy) to 7 (very happy); and recommendation of Game of Stones to others using the question: “On the basis of your experience of Game of Stones, how likely are you to recommend Game of Stones to other men?” with responses on a 7-point scale ranging from 1 (very unlikely) to 7 (very likely).

### Analysis

The analysis of the primary outcome estimated the mean difference in change from baseline weight expressed as a percentage at 24 months from baseline between 12 months text messaging with financial incentives compared to control; and 12 months text messaging alone compared to control, using a linear regression model adjusting for recruitment centre and method of recruitment route (GP or community) as a fixed effect. Secondary outcome measures were analysed using a variety of generalised linear models, including binary logit regression for dichotomous outcomes (e.g. number of participants achieving any weight loss) and ordered logit for ordinal outcomes (e.g. categorised weight loss). Statistical significance is at the α=2.5% level, consistent with the assumptions made in the 12-month sample size calculation. More information pertaining to the strategy used for each secondary outcome is specified in the associated statistical analysis plan^10^. The Number Needed to Treat (NNT) was derived from the relevant dichotomous outcomes If the NNT confidence interval includes a value of 0 or infinity this implies that the observed treatment effect is not statistically significant.

Analyses of the primary and all relevant secondary outcomes accounted for missing weight observations at 24 months using multiple imputation by chained equations. Specifically, using the predictive mean matching approach, with K nearest neighbours equal to 5 and the number of iterations equating to the percentage of participants with a missing primary outcome (∼36%). Sensitivity analyses were conducted for all observed cases excluding those individuals who indicated that they took weight loss pills, injections, or meal replacements: 1) between baseline and 24M or 2) between 12M and 24M. These analyses similarly applied predictive mean matching to impute these excluded (“missing”) values.

### Patient and Public Involvement

Game of Stones included Public and patient involvement (PPI) from inception and throughout all its conduct. Continuous and responsive public involvement are outlined in detail in reports of the feasibility study^14^ the trial protocol^9^ and will be published for the full trial^14^.

### Results

For 585 participants randomised, weight measurement at 24 months was completed for 377 participants (64.4%): 129 (65.8%) in the text messaging with financial incentives group, 114 (58.8%) in the text messaging alone group and 134 (68.7) in the control group (Figure 1). Most weight assessments (n=331; 87.8%) were in person on Game of Stones research scales within 23 days of target date; blinding to group allocation was retained for 348 participants (92.3%). See Appendix A for details on weight assessment methods for participants at 24 months.

**Figure 1:**
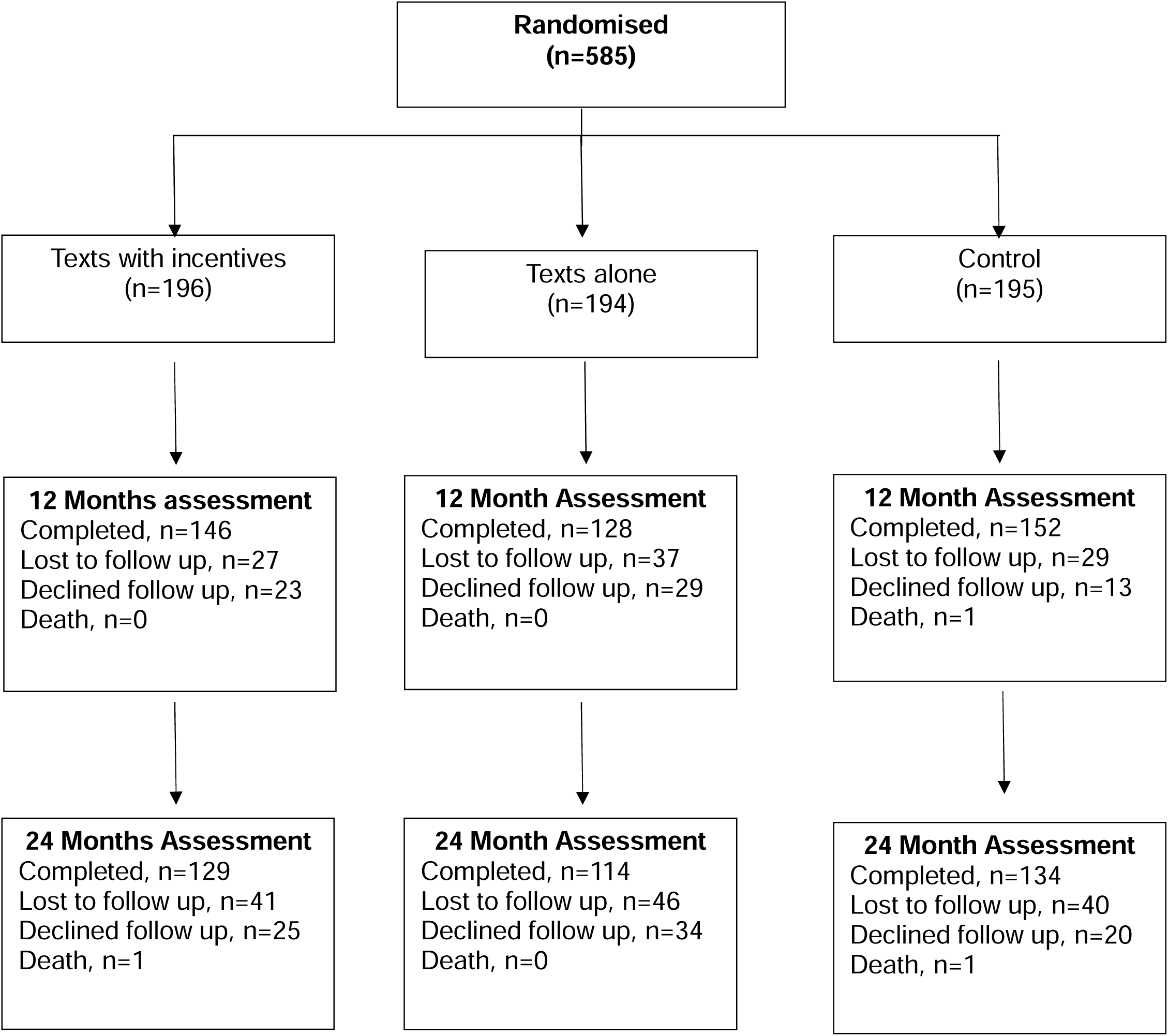
Participant groups and attendance at 12- and 24-month assessments. For additional details assessments before 12 months see^10^

Key baseline characteristics for participants completing the 24-month assessment are reported in Table 1. Baseline characteristics were comparable across trial groups. Information on all assessed baseline characteristics and how they were measured are reported elsewhere^10^. Participants had a mean BMI of 37.2kg/m^2^ (standard deviation (SD), 5.6) and a mean age of 51.3 (SD, 13.5) years. Most were of white ethnicity (91.0%) and reported one or more co-morbidities (72.4%). Underserved groups at 24 months included 36.6% men living in the two more deprived areas, 40.8% with multiple long-term conditions, 31.3% with a disability and 26.5% who had a diagnosed mental health condition. Baseline characteristics are broadly similar for participants providing a primary outcome at 24 months compared to those not providing an outcome. However, participants with weight data at 24 months had a lower mean weight at baseline than those for whom weight data are missing for the text messaging alone group (weight known: mean 114.9kg; SD, 16.0; n=114 compared to not provided mean 120.5kg; SD=19.9; n=80) and the waiting list control group (weight known: mean 115.5kg; SD, 21.3; n=134 compared to not provided: mean123.6kg; SD, 21.3; n=61). See Appendix B for details on baseline characteristics for participants with and without primary outcome information.

**Table 1:** Baseline characteristics for participants providing 24 months primary outcome.

|  | **Texts + Incentive**<br>**(N=129)** | **Texts Alone**<br>**(N=114)** | **Control**<br>**(N=134)** | **Total**<br>**(N=377)** |
| --- | --- | --- | --- | --- |
| **Age\* - mean (SD); n** | 51.4 (12.8); 129 | 53.0 (13.4); 114 | 49.7 (14.1); 134 | 51.3 (13.5); 377 |
| **Weight and Body Mass Index (BMI) - mean (SD)** |  |  |  |  |
| BMI (kg/m<sup>2</sup>) - mean (SD); n | 37.9 (5.9); 129 | 36.7 (4.2); 114 | 36.9 (6.2); 134 | 37.2 (5.6); 377 |
| ≥30-<35; n (%) | 49 (38.0) | 45 (39.5) | 66 (49.3) | 160 (42.4) |
| ≥35-<40; n (%) | 43 (33.3) | 44 (38.6) | 41 (30.6) | 128 (34.0) |
| ≥40; n (%) | 37 (28.7) | 25 (21.9) | 27 (20.1) | 89 (23.6) |
| **Deprivation Category - n (%)\*\*** |  |  |  |  |
| Most deprived areas | 32 (24.8) | 20 (17.5) | 29 (21.6) | 81 (21.5) |
| More deprived areas | 16 (12.4) | 23 (20.2) | 18 (13.4) | 57 (15.1) |
| Deprived areas | 19 (14.7) | 20 (17.5) | 23 (17.2) | 62 (16.4) |
| Less deprived areas | 23 (17.8) | 22 (19.3) | 18 (13.4) | 63 (16.7) |
| Least deprived areas | 39 (30.2) | 28 (24.6) | 45 (33.6) | 112 (29.7) |
| **Ethnic Group\* - n (%)** |  |  |  |  |
| Asian/ Asian British | 2 (1.6) | 1 (0.9) | 2 (1.5) | 5 (1.3) |
| Black/ African/ Caribbean/ Black British | 3 (2.3) | 2 (1.8) | 1 (0.7) | 6 (1.6) |
| Mixed/ multiple ethnic groups | 1 (0.8) | - | 3 (2.2) | 4 (1.1) |
| White | 118 (91.5) | 102 (89.5) | 123 (91.8) | 343 (91.0) |
| Other | 2 (1.6) | 2 (1.8) | 1 (0.7) | 5 (1.3) |
| Prefer not to say | 1 (0.8) | 3 (2.6) | - | 4 (1.1) |
| **Comorbidities\* - n (%)** |  |  |  |  |
| None | 29 (22.5) | 27 (23.7) | 31 (23.1) | 87 (23.1) |
| One or more | 90 (69.8) | 86 (75.4) | 97 (72.4) | 273 (72.4) |
| Multiple long-term conditions | 59 (45.7) | 52 (45.6) | 43 (32.1) | 154 (40.8) |
| Mental health condition | 34 (26.4) | 33 (28.9) | 33 (24.6) | 100 (26.5) |
| **Physical or Mental Disability\*\*\*** |  |  |  |  |
| Disability - n (%) | 44 (34.1) | 32 (28.1) | 42 (31.3) | 118 (31.3) |
| **Highest educational qualification\* - n (%)** |  |  |  |  |
| Degree level or above | 66 (51.2) | 44 (38.6) | 64 (47.8) | 174 (46.2) |
| Another kind of qualification | 57 (44.2) | 57 (50.0) | 59 (44.0) | 173 (45.9) |

\* Self-reported, \*\* To measure disadvantage participant postcodes of residence were looked up in the country-specific databases and assigned an index of multiple deprivation quintile. \*\*\* Disability was measured using the Office for National Statistics measures (see Hoddinott et al. (2024)<sup>10</sup> for details).

### Primary Outcomes

At 24-months, the mean percent weight change was −3.9% (SD, 6.9) for the text messaging with financial incentives group, −2.6% (SD, 6.8) for the text messaging alone group, and −2.2% (SD, 6.8) for the control group (Figure 2; Table 2). Compared with the control group, the text messaging with incentive group had greater weight loss which was not statistically significant (mean difference in percentage change from baseline, −1.0%; 97.5% CI, −2.6 to 0.6; P = .22). The text messaging alone and control groups had comparable weight loss (mean difference in percentage change from baseline, −0.0%; 97.5% CI, −1.6 to 1.5; P = .95). The pattern of results is similar for sensitivity analyses including only the 332 participants with verified weight assessments or including only the 302 participants not taking weight loss medication or meal replacements (Table 2, see Appendix C for full details of weight outcomes).

**Figure 2:**
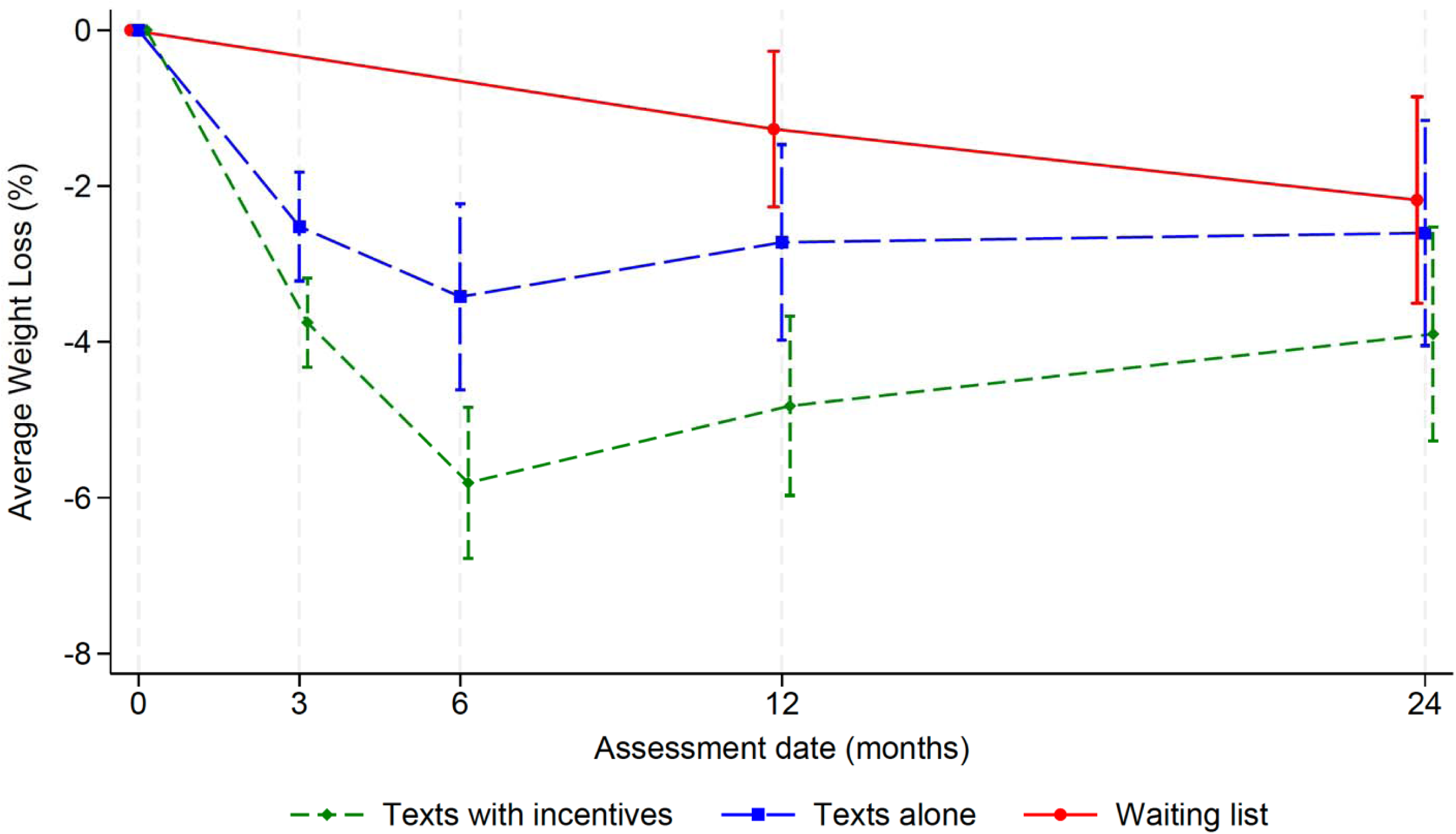
Weight loss over 24 months for trial groups. Values represent % mean weight loss (standard deviation).

**Table 2:** Primary and secondary weight outcomes at 24 months.

|  | Texts with<br>incentives | Texts alone | Waiting list<br>control | Mean difference<br>(Texts with Incentives<br>vs Control) 97.5% CI,<br>P-value | Mean difference<br>(Texts alone vs<br>Control) 97.5% CI, P-value |
| --- | --- | --- | --- | --- | --- |
| **Primary outcome** |  |  |  |  |  |
| % weight change - mean (SD); n |  |  |  |  |  |
| Observed Cases | -3.9 (6.9); 129 | -2.6 (6.8); 114 | -2.2 (6.8); 134 | -1.0 (-2.6, 0.6); 0.216* | -0.0 (-1.6, 1.5); 0.951* |
| Per protocol analysis** | -3.9 (7.1); 111 | -2.6 (6.5); 107 | -2.0 (7.0); 114 | -1.3 (-2.9, 0.4); 0.128* | -0.3 (-1.9, 1.4); 0.759* |
| Excluding WL med*** | -4.0 (7.1); 102 | -2.7 (6.9); 92 | -2.4 (6.9); 108 | -1.2 (-3.0, 0.5); 0.151* | -0.0 (-1.8, 1.7); 0.968* |
| BOCF | -2.6 (5.9); 196 | -1.5 (5.4); 194 | -1.5 (5.7); 195 | -1.1 (-2.3, 0.2); 0.067 | -0.0 (-1.3, 1.3); 0.984 |
| LOCF | -3.1 (6.0); 196 | -1.8 (5.7); 194 | -1.5 (5.9); 195 | -1.6 (-2.9, -0.2); 0.009 | -0.3 (-1.6, 1.1); 0.669 |
| **Secondary outcomes** |  |  |  |  |  |
| weight change in kg - mean (SD); n |  |  |  |  |  |
| Observed Cases | -4.7 (8.9); 129 | -2.8 (8.0); 114 | -2.7 (8.4); 134 |  |  |
| Per protocol analysis** | -4.6 (9.3); 110 | -2.9 (7.6); 101 | -2.6 (8.5); 112 |  |  |
| Excluding WL med*** | -4.9 (9.0); 102 | -2.9 (8.2); 92 | -2.8 (8.5); 108 |  |  |
| BOCF | -3.1 (7.6); 196 | -1.7 (6.3); 194 | -1.9 (7.0); 195 |  |  |
| LOCF | -3.8 (7.7); 196 | -2.0 (6.7); 194 | -1.9 (7.2); 195 |  |  |
| Weight loss categories (observed cases); n (%) |  |  |  |  |  |
| Weight gain | 32 (24.8) | 39 (34.2) | 46 (34.3) |  |  |
| 0<5% weight loss | 45 (34.9) | 43 (37.7) | 45 (33.6) |  |  |
| ≥5-<10% weight loss | 30 (23.3) | 22 (19.3) | 27 (20.1) |  |  |
| ≥10% weight loss | 22 (17.1) | 10 (8.8) | 16 (11.9) |  |  |

Note. \* Mean differences examined using Multiple Imputation by chained equations \*\* Per protocol analysis = assessment verified within 23 days on study scales, \*\* Excluding WL med = Excluding participants taking weight loss medications or meal replacements at any time from randomisation, BOCF = Baseline observation carried forward, LOCF = last observation carried forward, CI = Confidence Interval.

Weight loss outcomes by deprivation status at 12 and 24 months across the three trial groups are reported in see Appendix D. For the text messaging with financial incentives group the highest level of weight loss at 24 months was observed in the most deprived group (-5.5%; SD, 8.6; n= 32). Weight loss for the other deprivation categories was lower and variable. For the text messaging alone group the highest level of weight loss at 24 months was observed in the least deprived group (-4.4 %; SD, 8.5; n=28), with similar lower levels of weight loss across other deprivation categories.

### Secondary outcomes

At 24 months, participants had lost a mean of 4.7kg (SD, 8.9) in the text messaging with financial incentives group, 2.8kg (SD, 8.0) in the text messaging alone group, and 2.7kg (SD, 8.4) in the control group (Table 2). At 24 months, 97 of 129 participants (75.2%) in the text messaging with financial incentives group had lost some weight since baseline) compared with 75 of 114 (65.8%) in the text messaging alone group, and 88 of 134 (65.7%) in the control group (Table 2). At least 5% weight loss at 24 months was attained by 52 of 129 participants (40.3%) in the text messaging with financial incentives group compared with 32 of 114 (28.1%) in the text messaging alone group and 43 of 134 (32.1%) in the waiting list control group. At least 10% weight loss at 24 months was attained by 22 of 129 (17.1%) in the text messaging with financial incentives group, 10 of 114 (8.8%) in the text messaging alone group, and 16 of 134 (11.9%) in the control group. The number needed to treat (NNT) (97.5% CI) with text messaging with financial incentives for the outcome of weight loss of 5% or more at 24 months was 12 (NNT (harm) 22 to infinity to NNT (benefit) 5). For the outcome of weight loss of 10% or more at 24 months the NNT was 19 (NNT (harm) 25 to infinity to NNT (benefit) 7). For further details on secondary weight outcomes see Appendix E.

There were no statistically significant differences in PHQ-4, EQ-5D-5L; EQ-5D-5L visual analogue scale and EQ-5D-5L-AD scores between the text messaging with financial incentives group and the control group or between the text messaging alone group and the waiting list control group (see Table 3).

**Table 3:** Secondary outcomes of PHQ-4, EQ5D, EQ5D Visual Analog Scale and EQ-5D Anxiety/Depression at baseline and 24 months.

|  | Baseline |  |  | 24 months |  |  | Mean difference<br>(Texts with<br>Incentives<br>vs Control)<br>97.5% CI | Mean difference<br>(Texts alone<br>vs Control)<br>97.5% CI |
| --- | --- | --- | --- | --- | --- | --- | --- | --- |
|  | Texts with<br>Incentives | Texts<br>alone | Control | Texts with<br>Incentives | Texts<br>alone | Control |  |  |
| PHQ-4, mean (SD); n | 2.3 (2.5);<br>140 | 2.6 (2.7);<br>140 | 2.1 (2.3);<br>142 | 2.6 (3.1);<br>127 | 2.6 (3.0);<br>111 | 2.2 (2.4);<br>128 | 0.15<br>(-0.55,0.85) | 0.17<br>(-0.54,0.89) |
| EQ5D, mean (SD); n | 0.726<br>(0.229);<br>194 | 0.741<br>(0.230);<br>192 | 0.721<br>(0.207);<br>191 | 0.715<br>(0.266);<br>128 | 0.696<br>(0.266);<br>112 | 0.761<br>(0.219);<br>128 | -0.02<br>(-0.07,0.03) | -0.05<br>(-0.10,-0.00) |
| EQ5D Visual Analog<br>Scale, mean (SD); n | 61.6<br>(18.7);<br>194 | 62.9<br>(19.0);<br>186 | 62.3<br>(19.0);<br>193 | 66.9<br>(18.6);<br>125 | 66.0<br>(19.7);<br>111 | 68.0<br>(17.0);<br>125 | -0.17<br>(-5.01,4.68) | -1.50<br>(-6.53,3.52) |
| Anxiety/Depression<br>(EQ5D Dimension),<br>mean (SD); n | 1.8 (1.0);<br>194 | 1.8 (0.9);<br>193 | 1.9 (1.0);<br>194 | 1.9 (1.1);<br>128 | 1.9 (1.1);<br>114 | 1.8 (0.9);<br>129 | 0.18<br>(-0.04,0.40) | 0.18<br>(-0.04,0.41) |

Note. CI = Confidence Interval, EQ5D = EuroQol- 5 Dimension Questionnaire, PHQ-4 = Patient Health Questionnaire-4, SD = Standard deviation, n = number of participants.

For use of 13 assessed weight loss strategies, there were no statistically significant differences between the text messaging and financial incentives group compared to the control group and the text messaging alone group compared to the control group (see Appendix F).

Compared to the control group, significantly more participants would recommend the Game of Stones programme to others for both the text messaging and financial incentives group (mean difference at 24 months, 1.1%; 97.5% CI, 0.7 to 1.6) and text messaging alone group (mean difference at 24 months, 0.7%; 97.5% CI, 0.3 to 1.2). There were no differences in satisfaction with weight loss at 24 months between the text messaging and financial incentives group compared to the waiting list control group, or the text messaging alone group and the waiting list control group (see Appendix G).

## Discussion

The 24 month follow-up of the Game of Stones trial found weight loss favouring the text messaging with financial incentives group compared to the waiting list control group who received three-months of daily behavioural text-messages between months 12 to 15. The weight loss at 24 months for the text messaging alone group was similar to the waiting-list-control group. Participants in both intervention groups were significantly more likely to recommend Game of Stones to others compared to the waiting list control group.

The net weight loss between 12 and 24 months compares favourably to other studies. A systematic review of 155 behavioural weight management trials found an average weight change in intervention arms of –4.9 kg (SD, 3.8) at the end of the intervention (145 studies), and*–*4.0 kg (SD, 3.8) at 12 months after the intervention finished (47 studies)^15^. This level of weight change is similar to the text messaging with financial incentives group with -5.7 kg (SD 7.4) at 12 months^10^ and -4.7 kg (SD, 8.9) of weight loss at 24 months (12 months after the intervention ceased). In another systematic review of 67 behavioural weight management trials, pooled intervention groups compared to control groups lost 2.4 kg (95% CI, 2.8 to 1.9 kg) up to 18 months^16^. In this review, eight adults would need to enrol in a behavioural weight management programme for one to lose ≥5% of their starting weight by 12-18 months. The text messages with financial incentives group requires four men to enrol for one man to lose ≥5% of their starting weight by 12 months^10^ and 12 to enrol for one man to lose ≥5% at 24 months.

The socioeconomic patterning of weight loss for 24 months weight change suggests that Game of Stones is unlikely to contribute to increasing health inequalities^17^. Encouragingly, the highest level of weight loss at 24 months in the text messaging with financial incentives group was observed in participants living in the most deprived areas. The proportions of underserved groups recruited to the trial for area of deprivation, multiple long-term conditions, disability and mental health conditions were retained at 24 months.

The weight loss achieved in the current study is likely to be of clinical benefit^18^. A systematic review examining 124 behavioural weight management trials with a median follow up of 28 months suggests that any weight loss results in cardio-metabolic risk reduction which persist despite weight regain^19^.

Weight regain after behavioural interventions typically averages around 1-2 kg/year^20^. The weight regain in the text messaging with financial incentives group is within this range, and the text messaging alone group showed minimal average regain. A systematic review examining 249 behavioural weight management trials reports that financial incentives for weight loss are associated with faster weight regain at a rate of 1-1.5 kg/year compared to control conditions^21^. The current study does not suggest such a ‘backfiring effect’ of financial incentives in relation to weight regain. Although the expected levels of average weight regain in the text messaging with financial incentives group are higher compared to the text messaging alone group, this might relate to the higher levels of net weight loss, which are associated with higher levels of weight regain^21^.

The waiting list control group showed higher than expected weight changes at 12 and 24 months and continued to lose weight on average across the 24 months of the study. A systematic review of 29 control groups in behavioural weight management trials suggests that in the absence of a behavioural intervention, control group participants would weigh on average around 0.8 kg less than baseline at the end of the first year of follow-up^22^. The current waiting list control group weighed 1.5 kg (SD, 6.5) less at 12 months^10^. Another systematic review of 22 behavioural weight management trials found small net weight loss effects for control groups between 13-24 months (-0.20 kg, 95% CI − 0.49, 0.10, ≥ 12 months)^23^. These diminished longer-term weight effects in control groups compare with the findings of the current study, with the waiting list control group showing -2.7 (SD, 8.4) kg weight change at 24 months. The weight change pattern in the control group at 24 months is very likely related to the study design, where control group participants were offered behavioural texts for 3 months after the 12-month assessment (i.e. months 12-15). Weight loss in the control group of 2.7 kg at 24 months following three months of text messages, compares with 4.5 kg weight loss in the first three trial months in the text messaging alone group^10^. The greater weight loss for the text messaging alone group may be explained by the 3-month in person weight assessment effect, or the proximity to randomisation or the Covid-19 pandemic context.

The current study has several strengths. This is one of the first 24-month follow-up studies of either a text messaging or a financial incentive weight management trial. The interventions are pragmatic, scalable and low-burden for participants, providing an attractive option for weight management in resource restricted contexts. The study had an acceptable follow-up rate of 64% when considering the population who in large parts experience existing social, health or economic disadvantage.

This study has some limitations. The trial was designed to examine outcomes at 12 months, not 24 months. Consequently, the comparison between the interventions and the waiting list control group should be interpreted with caution and this follow-up should be seen as descriptive, rather than confirmatory. The waiting list control group received three months of behavioural text messages minimising its function as a typical control group in the context of the 24 month follow-up. Some imbalances in terms of baseline average weight for the text messaging alone and waiting list control groups were found, with participants providing a weight measurements at 24 months tending to be less heavy compared to those not followed up.

### Conclusion

At 24 months, participants receiving 12 months of behavioural text messages with financial incentives displayed continued weight loss compared to baseline with small regain following intervention end. The level of maintained weight loss is likely to be clinically important for some men. The results can inform longer-term health economic modelling of effects. The text messages alone showed similarly small average 24-month weight loss to the waiting list control group. This digital, scalable, low risk text messaging with financial incentives intervention has low participant and staff burden and shows promise for longer-term weight maintenance in men, including those living in the most deprived areas.

## Data Availability

All data produced in the present study are available upon reasonable request to the authors

## Notes

### Competing Interest Statement

Dr Hoddinott reported receiving grants from National Institute for Health Research (NIHR), and the Chief Scientist Office, Scotland, during the conduct of the study and serving as chair or member of Independent Trial Steering Committees unrelated to weight management trials; being a member of the NIHR School for Primary Care Research Funding panel. Dr Dombrowski reported receiving grants from the NIHR during the conduct of the study. Mr Swingler reported receiving grants from NIHR during the conduct of the study. Dr Cotton reported receiving grants from NIHR HTA (grant funding to institution) during the conduct of the study. Dr Avenell reported receiving grants from National Institute for Health and Care Research funding project in submission during the conduct of the study. Dr Hunt reported receiving grants from NIHR, the Australian Heart Foundation, and the Department of Health, Australia, during the conduct of the study; and serving as chair of the Health Improvement, Protection and Services Committee of the Chief Scientist Office. Dr MacLean reported receiving grants from the NIHR during the conduct of the study. Dr McKinley reported receiving grants from during the conduct of the study.Ms Torrens reported receiving grants from NIHR during the conduct of the study. Dr Turner reported receiving grants from NIHR during the conduct of the study and having had served as a member of the NIHR Health Technology Assessment (HTA) commissioning board, December 2017 to September 2020. Dr van der Pol reported receiving grants from NIHR Public Health Research and the Chief Scientist Office, Scotland, during the conduct of the study. Mr MacLennan reported receiving grants from the NIHR during the conduct of the study. No other disclosures were reported.

### Clinical Trial

ISRCTN91974895

### Funding Statement

This project was funded by the National Institute for Health and Care Research Public Health Research programme (NIHR PHR 129703)

### Author Declarations

Ethics committee North of Scotland Research Ethics Committee 2 gave ethical approval for this work [20/NS/0141].

