## Supplementary file for "Long-term Effects of Text Messages with Financial Incentives for Men with Obesity: Two-year Follow-up of the Game of Stones trial"

**Appendix A:** Weight assessment methods for men retained in the trial at 24 months

| | Texts with Incentives (N=129) | Texts alone (N=114) | Control (N=134) | Total (N=377) |
|---|---|---|---|---|
| Weight assessment in person on Game of Stones research scales within 23 days of target date | 111 (86.0) | 107 (93.9) | 113 (84.3) | 331 (87.8) |
| Weight assessment in person on Game of Stones research scales outside 23 days of target date | 4 (3.1) | 2 (1.8) | 3 (2.2) | 9 (2.4) |
| Blind to group allocation | 121 (93.8) | 107 (93.9) | 120 (89.6) | 348 (92.3) |
| Weighed on Game of Stones research scales remotely within 23 days of target date | - | - | 1 (0.7) | 1 (0.3) |
| Self-report weight within 23 days of target date | 10 (7.8) | 3 (2.6) | 15 (11.2) | 28 (7.4) |
| Self-report weight outside 23 days of target date | 4 (3.1) | 2 (1.8) | 2 (1.5) | 8 (2.1) |

**Appendix B:** Baseline characteristics by provision of primary outcome and treatment

| | Texts with Incentives Provided Outcome (N=129) | Texts with Incentives No Provided Outcome (N=67) | Texts Alone Provided Outcome (N=114) | Texts Alone No Provided Outcome (N=80) | Waiting List Control Provided Outcome (N=134) | Waiting List Control No Provided Outcome (N=61) |
|---|---|---|---|---|---|---|
| **Age* - mean (SD); n** | 51.4 (12.8); 129 | 47.4 (12.2); 66 | 53.0 (13.4); 114 | 50.0 (13.0); 80 | 49.7 (14.1); 134 | 51.1 (13.6); 61 |
| ≥18-<25 | 1 (0.8) | 1 (1.5) | 1 (0.9) | . (.) | 2 (1.5) | 1 (1.6) |
| ≥25-<45 | 35 (27.1) | 29 (43.3) | 32 (28.1) | 27 (33.8) | 50 (37.3) | 20 (32.8) |
| ≥45-<65 | 72 (55.8) | 32 (47.8) | 59 (51.8) | 40 (50.0) | 60 (44.8) | 30 (49.2) |
| ≥65-<75 | 18 (14.0) | 3 (4.5) | 16 (14.0) | 11 (13.8) | 16 (11.9) | 9 (14.8) |
| ≥75 | 3 (2.3) | 1 (1.5) | 6 (5.3) | 2 (2.5) | 6 (4.5) | 1 (1.6) |
| **Deprivation Category - n (%)** | | | | | | |
| Most deprived | 32 (24.8) | 16 (23.9) | 20 (17.5) | 16 (20.0) | 29 (21.6) | 21 (34.4) |
| More deprived | 16 (12.4) | 12 (17.9) | 23 (20.2) | 14 (17.5) | 18 (13.4) | 10 (16.4) |
| Deprived | 19 (14.7) | 6 (9.0) | 20 (17.5) | 13 (16.2) | 23 (17.2) | 6 (9.8) |
| Less deprived | 23 (17.8) | 16 (23.9) | 22 (19.3) | 18 (22.5) | 18 (13.4) | 13 (21.3) |
| Least deprived | 39 (30.2) | 16 (23.9) | 28 (24.6) | 18 (22.5) | 45 (33.6) | 11 (18.0) |
| **Ethnic Group* - n (%)** | | | | | | |
| White | 118 (91.5) | 61 (91.0) | 102 (89.5) | 72 (90.0) | 123 (91.8) | 49 (80.3) |
| Mixed/ multiple ethnic groups | 1 (0.8) | 1 (1.5) | . (.) | . (.) | 3 (2.2) | 1 (1.6) |
| Asian/ Asian British | 2 (1.6) | - | 1 (0.9) | - | 2 (1.5) | - |
| Black/ African/ Caribbean/ Black British | 3 (2.3) | - | 2 (1.8) | - | 1 (0.7) | - |
| Other | 2 (1.6) | 1 (1.5) | 2 (1.8) | 1 (1.2) | 1 (0.7) | 1 (1.6) |
| Prefer not to say | 1 (0.8) | - | 3 (2.6) | - | . (.) | - |
| **Relationship status* - n (%)** | | | | | | |

| | Texts with Incentives Provided Outcome (N=129) | Texts with Incentives No Provided Outcome (N=67) | Texts Alone Provided Outcome (N=114) | Texts Alone No Provided Outcome (N=80) | Waiting List Control Provided Outcome (N=134) | Waiting List Control No Provided Outcome (N=61) |
|---|---|---|---|---|---|---|
| Single (never married; never in a civil partnership) | 20 (15.5) | 10 (14.9) | 13 (11.4) | 6 (7.5) | 18 (13.4) | 9 (14.8) |
| Co-habiting | 16 (12.4) | 9 (13.4) | 17 (14.9) | 17 (21.2) | 25 (18.7) | 12 (19.7) |
| Married / civil partnership | 85 (65.9) | 41 (61.2) | 72 (63.2) | 44 (55.0) | 78 (58.2) | 35 (57.4) |
| Separated | 2 (1.6) | 3 (4.5) | 3 (2.6) | 3 (3.8) | 2 (1.5) | 1 (1.6) |
| Divorced | 4 (3.1) | 1 (1.5) | 4 (3.5) | 4 (5.0) | 5 (3.7) | 1 (1.6) |
| Widowed | - | - | - | - | - | - |
| Prefer not to say | 2 (1.6) | - | 2 (1.8) | - | . (.) | - |
| **Comorbidities* - n (%)** | | | | | | |
| None | 29 (22.5) | 16 (23.9) | 27 (23.7) | 25 (31.2) | 31 (23.1) | 12 (19.7) |
| One or more | 90 (69.8) | 46 (68.7) | 86 (75.4) | 50 (62.5) | 97 (72.4) | 47 (77.0) |
| Stroke (including TIA) | 7 (5.4) | 2 (3.0) | 2 (1.8) | 1 (1.2) | 6 (4.5) | 2 (3.3) |
| High Blood Pressure | 60 (46.5) | 33 (49.3) | 50 (43.9) | 33 (41.2) | 53 (39.6) | 33 (54.1) |
| Heart condition such as angina or atrial fibrillation | 19 (14.7) | 10 (14.9) | 25 (21.9) | 9 (11.2) | 17 (12.7) | 11 (18.0) |
| Diabetes | 27 (20.9) | 10 (14.9) | 22 (19.3) | 16 (20.0) | 13 (9.7) | 16 (26.2) |
| Cancer | 4 (3.1) | 2 (3.0) | 5 (4.4) | 3 (3.8) | 4 (3.0) | 1 (1.6) |
| Arthritis | 32 (24.8) | 8 (11.9) | 36 (31.6) | 19 (23.8) | 32 (23.9) | 15 (24.6) |
| Mental health condition | 34 (26.4) | 17 (25.4) | 33 (28.9) | 13 (16.2) | 33 (24.6) | 16 (26.2) |
| Possible Latent Mental Health Condition | 35 (27.1) | 15 (22.4) | 24 (21.1) | 24 (30.0) | 32 (23.9) | 12 (19.7) |

| | Texts with Incentives Provided Outcome (N=129) | Texts with Incentives No Provided Outcome (N=67) | Texts Alone Provided Outcome (N=114) | Texts Alone No Provided Outcome (N=80) | Waiting List Control Provided Outcome (N=134) | Waiting List Control No Provided Outcome (N=61) |
|---|---|---|---|---|---|---|
| Multiple Long-Term Conditions (MLTC) | 59 (45.7) | 23 (34.3) | 52 (45.6) | 30 (37.5) | 43 (32.1) | 28 (45.9) |
| MLTC including self-reported diabetes | 25 (19.4) | 8 (11.9) | 20 (17.5) | 14 (17.5) | 11 (8.2) | 12 (19.7) |
| **Physical or Mental Disability* - n (%)** | 59 (45.7) | 23 (34.3) | 45 (39.5) | 26 (32.5) | 57 (42.5) | 23 (37.7) |
| **Perceived wealth - mean (SD); n** | | | | | | |
| Perceives to live in relatively wealthy neighbourhood (0-100, Strongly disagree) | 56.9 (28.1); 128 | 54.3 (27.1); 63 | 55.4 (27.7); 107 | 54.1 (29.0); 70 | 56.2 (27.4); 125 | 53.5 (30.5); 57 |
| Feels relatively wealthy compared to others (0-100, Strongly disagree) | 54.8 (24.7); 128 | 52.6 (20.8); 63 | 52.3 (25.5); 105 | 49.1 (25.1); 69 | 54.0 (24.0); 122 | 52.7 (24.8); 57 |
| Feels like they have enough money (0-100, Strongly disagree) | 53.3 (29.7); 128 | 53.3 (26.3); 63 | 56.8 (29.4); 105 | 52.3 (29.4); 66 | 55.9 (27.8); 123 | 55.0 (26.9); 57 |
| **Financial Strain* - n (%)** | | | | | | |
| Living comfortably | 43 (33.3) | 20 (29.9) | 37 (32.5) | 21 (26.2) | 42 (31.3) | 16 (26.2) |
| Doing alright | 54 (41.9) | 30 (44.8) | 41 (36.0) | 30 (37.5) | 53 (39.6) | 27 (44.3) |
| Just about getting by | 18 (14.0) | 7 (10.4) | 23 (20.2) | 16 (20.0) | 19 (14.2) | 15 (24.6) |

| | Texts with Incentives Provided Outcome (N=129) | Texts with Incentives No Provided Outcome (N=67) | Texts Alone Provided Outcome (N=114) | Texts Alone No Provided Outcome (N=80) | Waiting List Control Provided Outcome (N=134) | Waiting List Control No Provided Outcome (N=61) |
|---|---|---|---|---|---|---|
| Finding it quite difficult | 8 (6.2) | 5 (7.5) | 6 (5.3) | 4 (5.0) | 10 (7.5) | 2 (3.3) |
| Finding it very difficult | 6 (4.7) | 1 (1.5) | 4 (3.5) | 1 (1.2) | 3 (2.2) | 1 (1.6) |
| Prefer not to say | - | 2 (3.0) | - | 4 (5.0) | - | . (.) |
| **Household composition* - n (%)** | | | | | | |
| Lives alone | 17 (13.2) | 8 (11.9) | 13 (11.4) | 8 (10.0) | 16 (11.9) | 6 (9.8) |
| Lives with partner | 99 (76.7) | 47 (70.1) | 91 (79.8) | 60 (75.0) | 103 (76.9) | 47 (77.0) |
| Lives with child/children | 59 (45.7) | 34 (50.7) | 46 (40.4) | 34 (42.5) | 46 (34.3) | 25 (41.0) |
| Lives with parents | 8 (6.2) | 5 (7.5) | 4 (3.5) | 3 (3.8) | 8 (6.0) | 9 (14.8) |
| Lives with friends | 2 (1.6) | - | 2 (1.8) | - | 1 (0.7) | - |
| Other | 6 (4.7) | 3 (4.5) | 2 (1.8) | 2 (2.5) | 1 (0.7) | 1 (1.6) |
| Household size - mean (SD); n | 2.8 (1.2); 128 | 2.9 (1.3); 64 | 2.6 (1.1); 112 | 2.9 (1.4); 78 | 2.5 (1.0); 127 | 2.8 (1.2); 60 |
| **Highest educational qualification* - n (%)** | | | | | | |
| Degree level or above | 66 (51.2) | 26 (38.8) | 44 (38.6) | 27 (33.8) | 64 (47.8) | 22 (36.1) |
| Another kind of qualification | 57 (44.2) | 33 (49.3) | 57 (50.0) | 38 (47.5) | 59 (44.0) | 29 (47.5) |
| **Employment Status* - n (%)** | | | | | | |

| | Texts with Incentives Provided Outcome (N=129) | Texts with Incentives No Provided Outcome (N=67) | Texts Alone Provided Outcome (N=114) | Texts Alone No Provided Outcome (N=80) | Waiting List Control Provided Outcome (N=134) | Waiting List Control No Provided Outcome (N=61) |
|---|---|---|---|---|---|---|
| Paid job - Full time (30+ hours per week) | 72 (55.8) | 48 (71.6) | 58 (50.9) | 42 (52.5) | 80 (59.7) | 34 (55.7) |
| Paid job - Part time (8-29 hours per week) | 7 (5.4) | 2 (3.0) | 7 (6.1) | 6 (7.5) | 6 (4.5) | 5 (8.2) |
| Paid job - Part time (Under 8 hours per week) | 1 (0.8) | - | 1 (0.9) | - | . (.) | - |
| Self-employed | 8 (6.2) | 6 (9.0) | 11 (9.6) | 10 (12.5) | 9 (6.7) | 4 (6.6) |
| Full time student | 2 (1.6) | - | 2 (1.8) | - | 1 (0.7) | - |
| Unemployed and seeking work | 2 (1.6) | - | . (.) | - | 2 (1.5) | - |
| Retired | 22 (17.1) | 4 (6.0) | 21 (18.4) | 14 (17.5) | 24 (17.9) | 13 (21.3) |
| Not in paid work due to illness or disability | 10 (7.8) | 1 (1.5) | 9 (7.9) | 1 (1.2) | 6 (4.5) | 2 (3.3) |
| Not in paid work for other reason | - | - | - | - | - | - |
| Other | 4 (3.1) | 2 (3.0) | . (.) | . (.) | 1 (0.7) | . (.) |
| Prefer not to say | - | 1 (1.5) | - | . (.) | - | . (.) |
| **Access to self-monitoring equipment* - n (%)** | | | | | | |
| Owns scales for self-weighing | 106 (82.2) | 60 (89.6) | 93 (81.6) | 63 (78.8) | 114 (85.1) | 51 (83.6) |
| Scales link to internet/app | 15 (11.6) | 14 (20.9) | 10 (8.8) | 6 (7.5) | 19 (14.2) | 12 (19.7) |

| | **Texts with Incentives Provided Outcome (N=129)** | **Texts with Incentives No Provided Outcome (N=67)** | **Texts Alone Provided Outcome (N=114)** | **Texts Alone No Provided Outcome (N=80)** | **Waiting List Control Provided Outcome (N=134)** | **Waiting List Control No Provided Outcome (N=61)** |
|---|---|---|---|---|---|---|
| Owns an activity tracker/pedometer | 65 (50.4) | 36 (53.7) | 59 (51.8) | 37 (46.2) | 88 (65.7) | 32 (52.5) |
| Highest weight (kg) - mean (SD); n | 125.7 (23.2); 126 | 126.7 (19.0); 64 | 120.6 (19.3); 109 | 128.0 (22.2); 76 | 121.9 (23.3); 133 | 129.0 (24.6); 58 |
| Lowest weight (kg) - mean (SD); n | 90.1 (18.9); 124 | 94.1 (18.4); 61 | 88.1 (15.9); 109 | 92.0 (15.8); 75 | 89.9 (16.0); 130 | 90.8 (20.5); 59 |
| Intended weight loss in study (kg) - mean (SD); n | 23.9 (17.8); 126 | 23.3 (12.1); 63 | 22.4 (15.1); 110 | 24.9 (16.2); 75 | 21.4 (15.4); 131 | 22.3 (10.3); 57 |
| Weight loss attempts - median (P25, P75); n | 7.0 (3.0-12.0); 123 | 6.0 (3.0-10.0); 65 | 6.0 (4.0-10.0); 110 | 5.0 (3.0-10.0); 77 | 5.0 (3.0-10.0); 133 | 5.0 (3.0-10.0); 59 |
| **Measured Weight and Height - n (%)** | | | | | | |
| Weight (kg) - mean (SD); n | 119.5 (21.0); 129 | 121.8 (18.1); 67 | 114.9 (16.0); 114 | 120.5 (19.9); 80 | 115.5 (21.3); 134 | 123.6 (21.3); 61 |
| Height (cm) - mean (SD); n | 177.4 (6.4); 129 | 177.9 (8.4); 67 | 176.7 (7.1); 114 | 177.7 (6.5); 80 | 176.8 (8.0); 134 | 176.8 (7.0); 61 |
| BMI (kg/m^2) - mean (SD); n | 37.9 (5.9); 129 | 38.6 (5.9); 67 | 36.7 (4.2); 114 | 38.1 (5.3); 80 | 36.9 (6.2); 134 | 39.6 (6.7); 61 |
| ≥30-<35; n (%) | 49 (38.0) | 17 (25.4) | 45 (39.5) | 28 (35.0) | 66 (49.3) | 16 (26.2) |
| ≥35-<40; n (%) | 43 (33.3) | 30 (44.8) | 44 (38.6) | 25 (31.2) | 41 (30.6) | 22 (36.1) |
| ≥40; n (%) | 37 (28.7) | 20 (29.9) | 25 (21.9) | 27 (33.8) | 27 (20.1) | 23 (37.7) |

* Self-reported

**Appendix C:** Percentage weight change and actual weight change (in kilograms) at 24 months from baseline (primary outcomes)

| **Weight Change (%) - mean (SD); n** | **Texts with Incentives (N=129)** | **Texts alone (N=114)** | **Control (N=134)** | **Mean Difference (Texts with Incentives vs Control) 97.5% CI, P- value** | **Mean Difference (Texts alone vs Control) 97.5% CI, P- value** |
|---|---|---|---|---|---|
| All Observed Cases | | | | | |
| Baseline Weight | 120.3 (20.1); 196 | 117.2 (17.9); 194 | 118.1 (21.6); 195 | | |
| 24 Month Weight | 114.8 (22.1); 129 | 112.1 (18.9); 114 | 112.8 (21.4); 134 | | |
| Actual Change in kg | -4.7 (8.9); 129 | -2.8 (8.0); 114 | -2.7 (8.4); 134 | | |
| Percentage Change | -3.9 (6.9); 129 | -2.6 (6.8); 114 | -2.2 (6.8); 134 | -1.7 (-3.6, 0.2); 0.045 | -0.4 (-2.4, 1.5); 0.617 |
| AOC_MICE | | | | -1.0 (-2.6, 0.6); 0.216 | -0.0 (-1.6, 1.5); 0.951 |
| Per Protocol 1 | | | | | |
| Baseline Weight | 120.3 (20.1); 196 | 117.2 (17.9); 194 | 118.1 (21.6); 195 | | |
| 24 Month Weight | 114.5 (20.3); 111 | 111.6 (18.1); 107 | 113.3 (21.8); 114 | | |
| Actual Change in kg | -4.7 (9.2); 111 | -2.9 (7.5); 107 | -2.5 (8.6); 114 | | |
| Percentage Change | -3.9 (7.1); 111 | -2.6 (6.5); 107 | -2.0 (7.0); 114 | -2.0 (-4.0, 0.1); 0.033 | -0.7 (-2.8, 1.4); 0.464 |
| PP1_MICE | | | | -1.3 (-2.9, 0.4); 0.128 | -0.3 (-1.9, 1.4); 0.759 |
| EWLM_AnyTime | | | | | |
| Baseline Weight | 120.3 (20.1); 196 | 117.2 (17.9); 194 | 118.1 (21.6); 195 | | |
| 24 Month Weight | 112.7 (20.2); 102 | 110.8 (18.7); 92 | 110.5 (19.6); 108 | | |
| Actual Change in kg | -4.9 (9.0); 102 | -2.9 (8.2); 92 | -2.8 (8.5); 108 | | |
| Percentage Change | -4.0 (7.1); 102 | -2.7 (6.9); 92 | -2.4 (6.9); 108 | -1.6 (-3.8, 0.6); 0.107 | -0.3 (-2.6, 1.9); 0.740 |
| MICE_EWLM_AnyTime | | | | -1.2 (-3.0, 0.5); 0.151 | -0.0 (-1.8, 1.7); 0.968 |

| | | | | | |
|---|---|---|---|---|---|
| EWLM_12M | | | | | |
| Baseline Weight | 120.3 (20.1); 196 | 117.2 (17.9); 194 | 118.1 (21.6); 195 | | |
| 24 Month Weight | 113.5 (21.7); 111 | 111.2 (18.3); 99 | 110.5 (19.5); 110 | | |
| Actual Change in kg | -4.8 (9.0); 111 | -2.7 (8.0); 99 | -2.8 (8.4); 110 | | |
| Percentage Change | -4.0 (7.1); 111 | -2.5 (6.7); 99 | -2.3 (6.9); 110 | -1.7 (-3.8, 0.5); 0.079 | -0.2 (-2.4, 1.9); 0.810 |
| MICE_EWLM_Last12M | | | | -1.6 (-3.5, 0.3); 0.107 | -0.3 (-2.3, 1.6); 0.740 |
| Baseline Observation Carried Forward | | | | | |
| Baseline Weight | 120.3 (20.1); 196 | 117.2 (17.9); 194 | 118.1 (21.6); 195 | | |
| 24 Month Weight | 117.2 (21.0); 196 | 115.6 (19.7); 194 | 116.2 (21.9); 195 | | |
| Actual Change in kg | -3.1 (7.6); 196 | -1.7 (6.3); 194 | -1.9 (7.0); 195 | | |
| Percentage Change | -2.6 (5.9); 196 | -1.5 (5.4); 194 | -1.5 (5.7); 195 | -1.1 (-2.3, 0.2); 0.067 | -0.0 (-1.3, 1.3); 0.984 |
| Last Observation Carried Forward | | | | | |
| Baseline Weight | 120.3 (20.1); 196 | 117.2 (17.9); 194 | 118.1 (21.6); 195 | | |
| 24 Month Weight | 116.5 (20.8); 196 | 115.3 (19.8); 194 | 116.2 (22.0); 195 | | |
| Actual Change in kg | -3.8 (7.7); 196 | -2.0 (6.7); 194 | -1.9 (7.2); 195 | | |
| Percentage Change | -3.1 (6.0); 196 | -1.8 (5.7); 194 | -1.5 (5.9); 195 | -1.6 (-2.9, -0.2); 0.009 | -0.3 (-1.6, 1.1); 0.669 |

Note. CI = confidence interval; kg = kilograms; M = months; n = number of participants; SD = standard deviation; AOC_MICE = All observed cases (using) Multiple Imputation by chained equations, PP1_MICE = per protocol 1 [Strict/gold-standard: assessment verified within 23 days on scales] (using) Multiple Imputation by chained equations; EWLM_AnyTime = Excluding participants taking weight loss medications at any time from randomisation; MICE_EWLM_AnyTime = Excluding participants taking weight loss medications taken at any time from randomisation (using) Multiple Imputation by chained equations; MICE_EWLM_Last12M = Excluding participants taking weight loss medications taken from 12M-24M post randomisation (using) Multiple Imputation by chained equations.

**Appendix D**: Weight loss percentage by deprivation status at 12 and 24 months from baseline for intervention and control groups.

| **Trial group** | **Deprivation status (IMD*)** | **12 Months Weight Loss (%) mean (SD); n** | **24 Months Weight Loss (%) mean (SD); n** |
|---|---|---|---|
| Texts + Incentive | 1 (most deprived) | -5.6 (7.0); 37 | -5.5 (8.6); 32 |
| | 2 | -5.7 (5.8); 19 | -3.4 (5.7); 16 |
| | 3 | -6.9 (6.6); 20 | -4.5 (7.3); 19 |
| | 4 | -2.7 (4.7); 24 | -2.0 (4.7); 23 |
| | 5 (least deprived) | -4.1 (5.8); 46 | -3.6 (6.7); 39 |
| Texts alone | 1 (most deprived) | -3.6 (4.4); 22 | -1.4 (4.8); 20 |
| | 2 | -3.1 (5.6); 25 | -1.8 (6.6); 23 |
| | 3 | -3.4 (8.6); 19 | -2.7 (8.0); 20 |
| | 4 | -1.0 (4.5); 29 | -2.1 (5.1); 22 |
| | 5 (least deprived) | -3.2 (7.6); 32 | -4.4 (8.5); 28 |
| Waiting List Control | 1 (most deprived) | -3.0 (6.0); 34 | -2.6 (7.3); 29 |
| | 2 | -1.3 (6.2); 22 | -2.4 (9.4); 18 |
| | 3 | -0.9 (4.6); 24 | -4.2 (5.9); 23 |
| | 4 | -1.2 (5.2); 22 | -2.8 (4.3); 18 |
| | 5 (least deprived) | -0.3 (5.1); 49 | -0.6 (6.5); 45 |

*Note*. n= number of participants, SD = standard deviation, *Index of Multiple Deprivation ranging 1 (most) to 5 (least).

**Appendix E:** Weight change at 24 months from baseline (secondary outcomes)

| | **Texts + Incentive** | **Texts alone** | **Control** | **Mean difference or odds ratio/risk difference (Texts + Incentive vs Control) 97.5% Confidence Interval** | **Mean difference or odds ratio/risk difference (Texts alone vs Control) 97.5% Confidence Interval** |
|---|---|---|---|---|---|
| Absolute Weight change in kg (any weight measurement - complete cases) - mean (SD); n | -4.7 (8.9); 129 | -2.8 (8.0); 114 | -2.7 (8.4); 134 | -1.8 (-4.2, 0.5) | -0.1 (-2.5, 2.3) |
| Weight loss dichotomies (any weight measurements - complete cases); n (%) | | | | | |
| >0% weight loss | 97 (75.2) | 75 (65.8) | 88 (65.7) | 1.6 (0.9, 2.9) | 1.0 (0.5, 1.8) |
| Multiple Imputation: >0% weight loss | | | | 1.3 (0.7, 2.3) | 1.0 (0.6, 1.7) |
| ≥5% | 52 (40.3) | 32 (28.1) | 43 (32.1) | 1.4 (0.8, 2.6) | 0.8 (0.4, 1.6) |
| Multiple Imputation: ≥5% weight loss | | | | 1.3 (0.8, 2.2) | 0.9 (0.5, 1.5) |
| ≥10% | 22 (17.1) | 10 (8.8) | 16 (11.9) | 1.6 (0.7, 3.5) | 0.7 (0.3, 1.9) |

| | | | | | |
|---|---|---|---|---|---|
| Multiple Imputation: >10% weight loss | | | | 1.3 (0.6, 2.6) | 0.8 (0.4, 1.7) |
| Weight change categories (any weight measurements - complete cases); n (%) | | | | 1.5 (0.9, 2.5) | 0.9 (0.5, 1.5) |
| Weight gain | 32 (24.8) | 39 (34.2) | 46 (34.3) | | |
| 0<5% weight loss | 45 (34.9) | 43 (37.7) | 45 (33.6) | | |
| ≥5-<10% weight loss | 30 (23.3) | 22 (19.3) | 27 (20.1) | | |
| ≥10% weight loss | 22 (17.1) | 10 (8.8) | 16 (11.9) | | |

**Appendix F:** Weight management strategies used at baseline and 24 Months

| | Texts + Incentive | Texts alone | Control | Texts + Incentive | Texts alone | Control | Odds Ratio (Texts + Incentive vs Control) 97.5% Confidence Interval | Odds Ratio (Texts alone vs Control) 97.5% Confidence Interval |
|---|---|---|---|---|---|---|---|---|
| Current Weight Management Strategies* | | | | | | | | |
| Kept track of weight by weighing yourself^ | | | | 100/129 (77.5) | 77/114 (67.5) | 95/133 (71.4) | 1.4 (0.7, 2.7) | 0.8 (0.4, 1.6) |
| Looked up strategies, tips, plans on how to lose weight | 106/195 (54.4) | 95/193 (49.2) | 106/195 (54.4) | 37/129 (28.7) | 39/114 (34.2) | 59/133 (44.4) | 0.5 (0.3, 0.9) | 0.7 (0.3, 1.2) |
| Avoided certain foods | 157/195 (80.5) | 147/193 (76.2) | 145/195 (74.4) | 83/129 (64.3) | 81/114 (71.1) | 91/133 (68.4) | 0.8 (0.4, 1.5) | 1.1 (0.6, 2.0) |
| Had a weight goal to work towards | 71/195 (36.4) | 76/193 (39.4) | 80/195 (41.0) | 65/129 (50.4) | 42/114 (36.8) | 74/133 (55.6) | 0.9 (0.5, 1.6) | 0.5 (0.3, 0.9) |
| Reminded yourself of the reasons you're trying to lose weight | 117/195 (60.0) | 120/193 (62.2) | 110/195 (56.4) | 90/129 (69.8) | 70/114 (61.4) | 87/133 (65.4) | 1.2 (0.7, 2.2) | 0.7 (0.4, 1.4) |
| Swapped one type of food for another | 98/195 (50.3) | 94/193 (48.7) | 100/195 (51.3) | 47/129 (36.4) | 55/114 (48.2) | 52/133 (39.1) | 0.9 (0.5, 1.7) | 1.6 (0.9, 2.9) |

| | | | | | | | | |
|---|---|---|---|---|---|---|---|---|
| Swapped one type of drink for another | 94/195 (48.2) | 90/193 (46.6) | 95/195 (48.7) | 47/129 (36.4) | 46/114 (40.4) | 49/133 (36.8) | 1.0 (0.5, 1.8) | 1.2 (0.7, 2.2) |
| Told others about your weight loss goals | 71/195 (36.4) | 72/193 (37.3) | 74/195 (37.9) | 52/129 (40.3) | 33/114 (28.9) | 51/133 (38.3) | 1.2 (0.6, 2.1) | 0.7 (0.4, 1.3) |
| Used a book, website, or app | 83/195 (42.6) | 78/193 (40.4) | 90/195 (46.2) | 33/129 (25.6) | 39/114 (34.2) | 54/133 (40.6) | 0.5 (0.3, 0.9) | 0.8 (0.4, 1.7) |
| Checked the portion size of things you eat | 94/195 (48.2) | 101/193 (52.3) | 108/195 (55.4) | 61/129 (47.3) | 63/114 (55.3) | 69/133 (51.9) | 0.8 (0.5, 1.4) | 1.2 (0.6, 2.1) |
| Kept track of the calorie/nutritional content of the things you eat and drink | 78/195 (40.0) | 78/193 (40.4) | 87/195 (44.6) | 42/129 (32.6) | 28/114 (24.6) | 53/133 (39.8) | 0.8 (0.4, 1.4) | 0.5 (0.3, 1.0) |
| Used a weight loss service to help me manage my weight | 31/195 (15.9) | 30/193 (15.5) | 49/195 (25.1) | 11/129 (8.5) | 14/114 (12.3) | 14/133 (10.5) | 1.1 (0.4, 3.1) | 2.1 (0.7, 6.2) |
| Cut down on alcohol | 75/195 (38.5) | 80/193 (41.5) | 87/195 (44.6) | 54/129 (41.9) | 46/114 (40.4) | 54/133 (40.6) | 1.2 (0.7, 2.2) | 1.0 (0.5, 1.8) |
| Increased the amount of physical activity, sport or exercise that you were doing | 136/195 (69.7) | 124/193 (64.2) | 129/195 (66.2) | 75/129 (58.1) | 69/114 (60.5) | 74/133 (55.6) | 1.1 (0.6, 2.0) | 1.2 (0.7, 2.2) |
| None | 10/195 (5.1) | 12/193 (6.2) | 9/195 (4.6) | 3/129 (2.3) | 2/114 (1.8) | 6/133 (4.5) | 0.3 (0.1, 2.0) | 0.3 (0.0, 2.4) |
| Another Strategy | 78/167 (46.7) | 56/160 (35.0) | 62/162 (38.3) | 35/110 (31.8) | 28/93 (30.1) | 32/110 (29.1) | 1.1 (0.5, 2.2) | 1.0 (0.5, 2.1) |

* Self-reported ^ Not collected at baseline

**Appendix G:** Participant recommendation of the Game of Stones trial and weight loss satisfaction at 24 months

| Variable - mean (SD); n | Texts with Incentives | Texts alone | Waiting List Control | Mean difference (Texts + Incentive vs Control) 97.5% Confidence Interval | Mean difference (Texts alone vs Control) 97.5% Confidence Interval |
|---|---|---|---|---|---|
| Programme recommendation* | 5.9 (1.5); 129 | 5.5 (1.5); 114 | 4.7 (1.7); 130 | 1.1 (0.7, 1.6) | 0.7 (0.3, 1.2) |
| Happy with weight loss progress* | 3.7 (1.7); 129 | 3.7 (1.6); 114 | 3.7 (1.5); 129 | 0.0 (-0.4, 0.5) | -0.0 (-0.5, 0.4) |

* Self-reported, both scores range 1-7
